# Machine-Learning Enhanced Diffusion Tensor Imaging with Four Encoding Directions

**DOI:** 10.1101/2024.08.19.24312228

**Authors:** Joshua Mawuli Ametepe, James Gholam, Leandro Beltrachini, Mara Cercignani, Derek Kenton Jones

## Abstract

**Purpose:** This study aims to reduce Diffusion Tensor MRI (DT-MRI) scan time by minimizing diffusion-weighted measurements. Using machine learning, DT-MRI parameters are accurately estimated with just four tetrahedrally-arranged diffusion-encoded measurements, instead of the usual six or more. This significantly shortens scan duration and is particularly useful in ultra-low field (ULF) MRI studies and for non-compliant populations (e.g., children, the elderly, or those with movement disorders) where long scan times are impractical.

**Methods:** To improve upon a previous tetrahedral encoding approach, this study used a deep learning (DL) model to predict parallel and radial diffusivities and the principal eigenvector of the diffusion tensor with four tetrahedrally-arranged diffusion-weighted measurements. Synthetic data were generated for model training, covering a range of diffusion tensors with uniformly distributed eigenvectors and eigenvalues. Separate DL models were trained to predict diffusivities and principal eigenvectors, then evaluated on a digital phantom and in vivo data collected at 64 mT.

**Results:** The DL models outperformed the previous tetrahedral encoding method in estimating diffusivities, fractional anisotropy, and principal eigenvectors, with significant improvements in ULF experiments, confirming the DL approach’s feasibility in low SNR scenarios. However, the models had limitations when the tensor’s principal eigenvector aligned with the scanner’s axes

**Conclusion:** The study demonstrates the potential of using DL to perform DT-MRI with only four directions in ULF environments, effectively reducing scan durations and addressing numerical instability seen in previous methods. These findings open new possibilities for ULF DT-MRI applications in research and clinical settings, particularly in pediatric neuroimaging

## 1.0 INTRODUCTION

Diffusion-weighted Imaging (DWI) is sensitive to the microscopic motion of water molecules within tissue, thus providing a unique contrast which complements T_1_ and T_2_-weighted MRI. As diffusion is anisotropic in white matter, typically 3 images with orthogonal diffusion directions are acquired and averaged to provide rotationally-invariant mean apparent diffusion coefficient (ADC) maps. Clinically, DWI has become the standard diagnostic scan for acute stroke and is routinely acquired in other conditions such as multiple sclerosis, brain tumours and dementia^1^. Diffusion anisotropy and tissue orientation can also be derived from DWI, within the framework of diffusion tensor MRI (DT-MRI) ^2,3^, which allows for the detailed visualization of tissue microstructure ^4^. As the diffusion tensor has six degrees of freedom, it requires a minimum of 6 diffusion-weighted images to estimate it unequivocally. On higher field systems, with accompanying higher SNR, and therefore less need to collect multiple averages of a signal, this requirement can be met within practical acquisition times. However, conducting DT-MRI at low and ultra-low fields (LF/ULF) presents significant challenges. DWI encodes the effects of diffusion as a signal attenuation, and therefore inherently results in low SNR images. This SNR problem is further exacerbated at low field ^5^. While increasing the number of averages can offset low SNR, this becomes an issue in non-compliant populations, like infants, children, or individuals who find it difficult to stay still for long periods.

To address this issue, this study re-visits the concept of tetrahedral encoding in DT-MRI, first introduced by Conturo et al.^6^, to assess its effectiveness at low field. This method hinges on the assumption of cylindrical symmetry of the diffusion tensor, allowing for a reduction in the minimum number of required measurements from six to four ^6^, effectively enabling the estimation of anisotropy and tissue orientation with just one extra measurement compared to the standard clinical three-direction ADC protocol.

We first explore the conceptual limitations of this standard tetrahedral encoding approach, including the challenges of estimating tensor components for certain tensor orientations. While one previous work ^7^ attempted to show the errors in fractional anisotropy (FA) estimates using tetrahedral encoding, it did not investigate the bias in tensor orientations, nor the origin of such errors. Here we provide a theoretical explanation for the bias. Next, using Monte Carlo simulations, we compare the noise sensitivity of the reduced tetrahedral encoding approach to that of a full six-direction encoding approach. We then leverage deep learning (DL) to address the challenges posed by low SNR and the limitations of tetrahedral encoding. By developing and validating DL models trained on synthetically generated data, we aim to reduce scan durations while preserving the integrity of DT-MRI results, even in the challenging low SNR context of LF/ULF MRI.

This paper is organized into five main sections: Theory; Methods; Results; Discussion, and Conclusion. The theory section provides a brief introduction to the tetrahedral encoding scheme as outlined by Conturo et al ^6^, offering a theoretical explanation of some issues we encountered with this method. The methods section describes our machine learning approach to address these issues and includes tests to compare our method with the standard tetrahedral approach. The results section presents the outcomes of these tests. Finally, the discussion and conclusion sections highlight the progress made, the limitations of our approach, and ideas for possible improvements to the method.

## 2.0 THEORY

Estimating a general diffusion tensor typically requires at least six diffusion-weighted images and one b=0 s/mm^2^ image^2,3^. However, Conturo and colleagues introduced the tetrahedral encoding scheme for DT-MRI, enabling tensor estimation with just four diffusion-weighted images under the assumption of cylindrical symmetry. In general, a diffusion tensor can be decomposed into three eigenvalues (λ_1_≤ λ_2_ ≤ λ_3_) and rotational angles (θ, ∅, Ψ)^6^. Assuming cylindrical symmetry (λ_2_ = λ_3_) simplifies the tensor to four variables: λ_1_, λ_2_, θ, and ∅, allowing us to estimate the tensor with only four diffusion-weighted measurements.

The success of this approach depends, in addition to the cylindrical symmetry assumption, on the even distribution of the four diffusion gradient orientations, achievable through a tetrahedral gradient configuration, which aligns the gradient orientations with the vertices of a regular tetrahedron with vertices

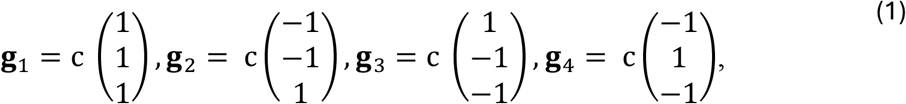

Where 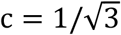.

The gradient vectors are visualised in figure 1. This simplification allows us to write the equation for the ADC along each encoding axis as

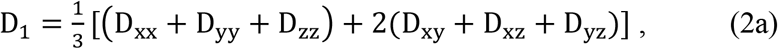

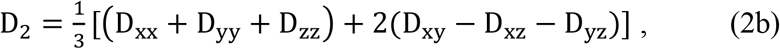

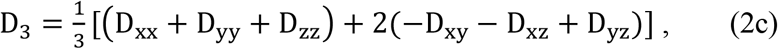

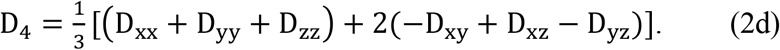

**Figure 1.**
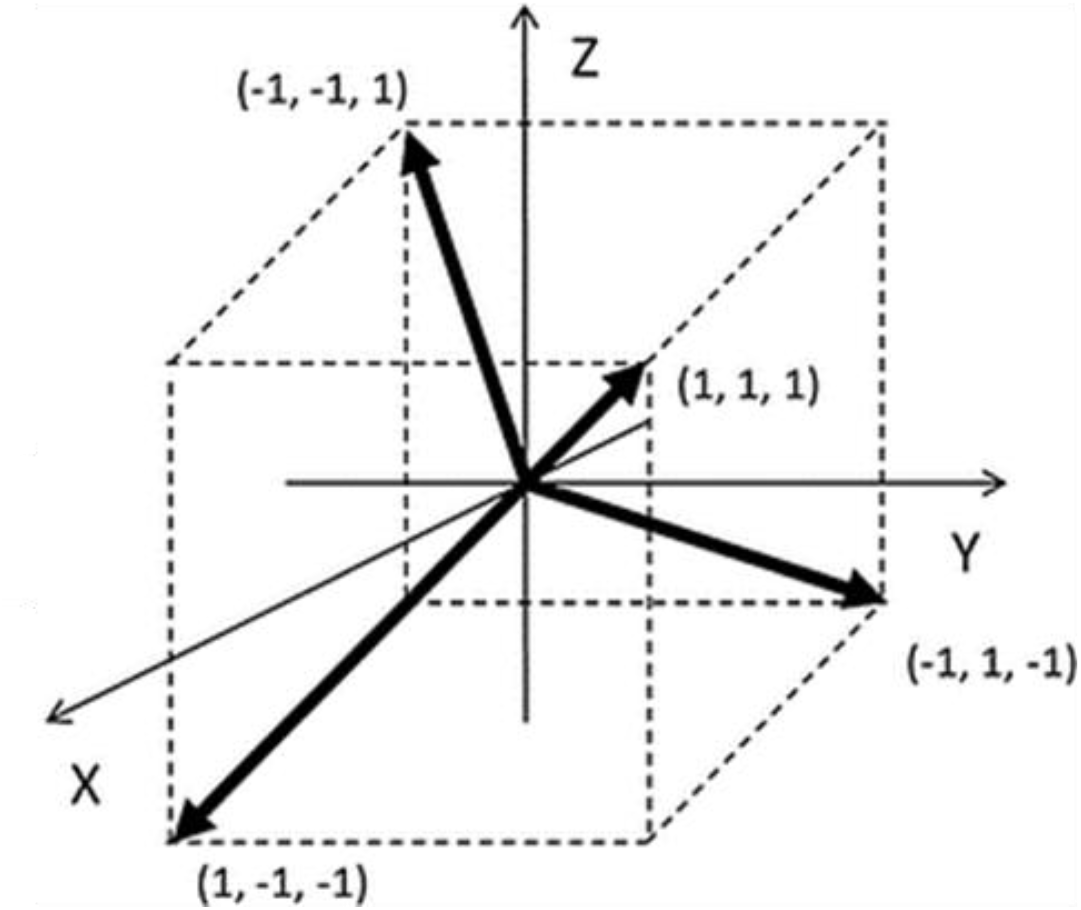
Tetrahedral gradient orientations employed in this work. The four gradient vectors are displayed in a 3d cartessian space pointing to the edges of a cube for visualisation. Image is taken from Ni et al, 2016 ^4^

It can be easily seen that the arithmetic mean of these four expressions is the mean diffusivity^6^, i.e.,

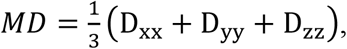

and it is then trivial to estimate the off-diagonal elements. Under the assumption of cylindrical symmetry, the principal eigenvector, parallel and radial diffusivities can be accurately estimated from the off-diagonal components using the following expressions ^6^,

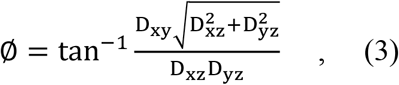

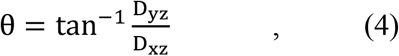

The parallel and radial diffusivities are given by

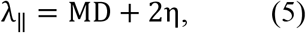

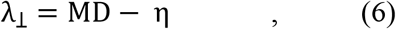

where

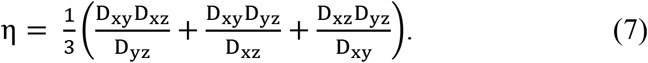

### 2.1 THE AXIS PROBLEM

A problem is encountered when a tensor’s eigenvectors are oriented along the ***x, y***, and ***z*** axes since the tensor’s off-diagonal components [i.e., the denominator in eqs. (3-7)] become zero. The closer the off-diagonal elements are to zero, the larger the error will be ^7^. Moreover, the standard tetrahedral approach to estimating the tensor hinges on differential signal attenuations across different gradient directions. Looking at eq. (2), when the tensor is rotated such that all off-diagonal elements are zero, each ADC is equal to the mean diffusivity, suggesting isotropy.

The same issue persists even when only the principal eigenvector aligns with the scanner’s axes. Under ideal circumstances, variations in the second and third eigenvalues would lead to differences in the measured signals. However, assuming cylindrical symmetry equates the second and third eigenvalues, rendering the orientations of the second and third eigenvectors inconsequential (see Figure 2). Consequently, when the tensor’s principal orientation aligns closely with any of the scanner’s axes, measurement variance decreases, resulting in a more isotropic appearance. This is a mathematical limitation of using only four measurements in DTI.

**Figure 2.**
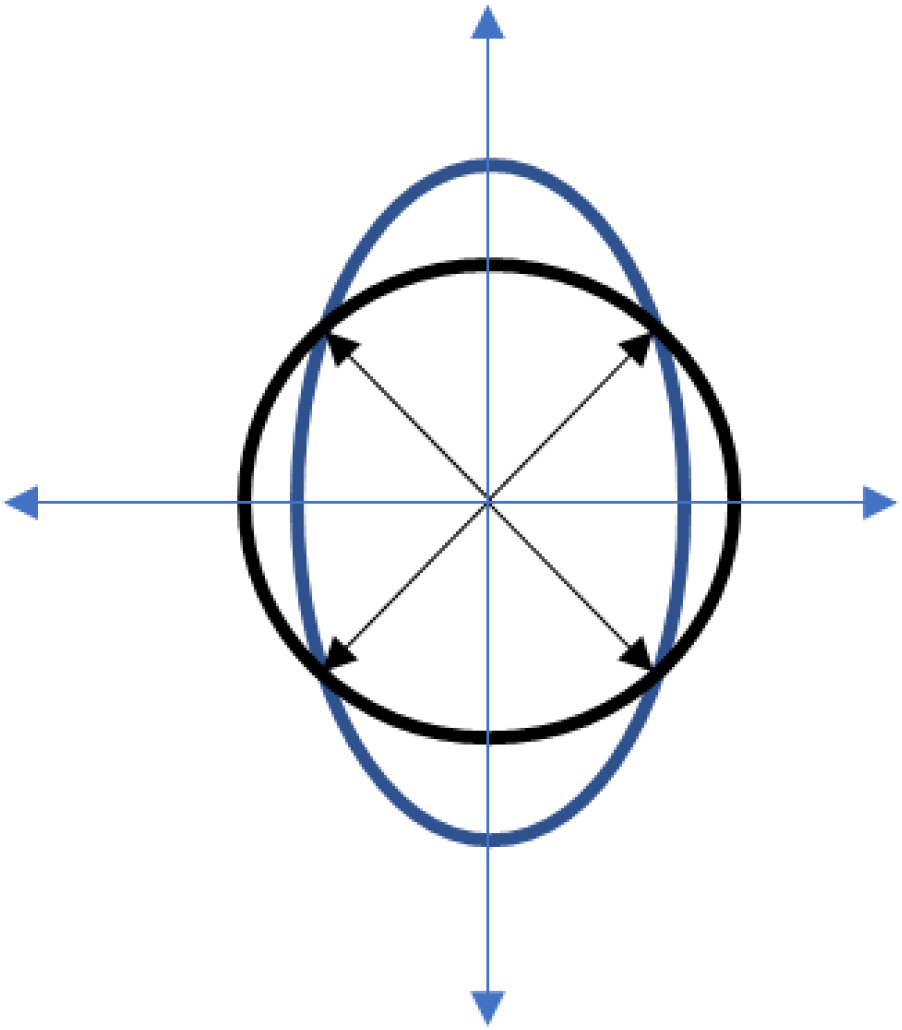
2D visual representation of the axis problem. The image shows an ellipse aligned with the vertical axes overlayed, with a circle, with gradient vectors at 45-degree angles extending from the centre to the edges. We see that the points at which the vectors touch the ellipse are the same points at which they touch the circle, showing that we can fit both an ellipse and a circle to the same measurements. This illustrates that with the tetrahedral gradient configuration, when the tensor aligns with the scanner’s axes, the measurements correspond to those of a sphere, demonstrating the isotropic appearance despite anisotropy in the actual data.

Finally, eq. (2) expresses the ADC as the mean diffusivity plus a function of the tensor’s off-diagonal components ^6^. This relationship is disrupted if the gradients deviate from a precise 45-degree orientation relative to the scanner’s axes. Participant motion during scanning requires corrective registration, involving affine transformations that rotate gradient vectors differently ^9^, complicating the underlying mathematics. Ignoring these gradient rotations introduces a significant limitation impacting the accuracy of our tensor estimations, particularly in real-world scenarios where patient movement is common.

## 3.0 METHODS

### 3.1 MACHINE LEARNING

#### 3.1.1 TRAINING DATA GENERATION

We developed a deep learning model to predict parallel and radial diffusivities (λ_1_ and λ_2_), as well as the principal eigenvector using four diffusion-weighted images. This required a representative dataset for training, which would typically be collected from a variety of brain samples. However, biases from data collection methods, scanner settings, noise, and potential hallucination artefacts can affect accuracy. To mitigate these biases, we used synthetic data for model training.

The first step involved generating tensors with uniformly distributed eigenvalues and angles of eigenvectors. The three eigenvectors were uniformly dispersed in three-dimensional space and eigenvalues were sampled within a predefined range for each tensor.

Our dataset comprised 14 million diffusion tensors. For each tensor, the angular parameters θ, ∅, and Ψ were uniformly sampled between 0 and 360 degrees. Eigenvalues were sampled within a range of 1.0 x 10^-4^ mm^2^/s to 3.0 x 10^-3^ mm^2^/s to avoid biases while covering expected voxel values ^8^. The sampled values were ordered as the first, second and third eigenvalues, with the mean of the second and third eigenvalues used to simulate a cylindrically symmetric tensor. Tetrahedral gradient vectors [see eq. (1)] were used to generate the diffusion-weighted signals.

To account for rotation issues due to registration, each dataset applied a randomly generated rotation matrix to the four gradient vectors. These matrices were generated by specifying rotation angles between 0 and 20 degrees in the ***x, y***, and ***z*** directions.

Using a b-value range of 500 s/mm^2^ to 1000 s/mm^2^, and a predetermined b_0_ signal value, we can derive our four diffusion-weighted signals. To simulate realistic conditions, Rician noise was added ^9^, based on a chosen SNR equal to 5 (typical for an image taken at 64 mT).

#### 3.1.2 FEATURE ENGINEERING

Organizing features effectively is crucial for our machine learning algorithm’s performance. Our input feature set included a b_0_ signal value, four DWI signal values, a b-value, and four gradient vectors, totalling 18 distinct features per sample, each with its own unique range. To promote model convergence and ensure fair consideration of all features, we adopted a standardized approach. We converted all inputs into vectors, framing the problem geometrically. These four vectors represent gradient directions, with their magnitudes indicating the Apparent Diffusion Coefficient (ADC). In our dataset, each of these vectors (i.e., ***g***_*i*_, *i* = 1, …, 4) corresponds to a gradient vector, where the vector length represents the ADC in its respective direction. So, for each gradient direction, the input vector is

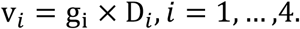

This approach streamlines our input features to 12, ensuring uniformity across our dataset. By reducing feature count, we enhanced model convergence and ensure that each feature contributes equally to the output.

We developed a multi-layer perceptron ^10^ trained on these standardised input data, creating two distinct models: one for predicting diffusivities, and another for estimating the principal eigenvector. The diffusivity model did not enforce the assumption of symmetry on the dataset, whereas it was enforced for the principal eigenvector model. Each perceptron has three layers preceding the output layer with Rectified linear Unit (ReLU) activation function which sets all negative outputs to zero. For the eigenvector model, the activation function in the third layer was set to tanh, which puts all outputs into a range of -1 and 1.

Training separate models allowed us to customize loss functions tailored specifically to diffusivities or vectors. Both models were trained on the noisy and noiseless versions of the data. For diffusivities, the mean squared error was chosen as the appropriate loss function suited to discrete and non-correlated scalar quantities. Conversely, predicting the principal eigenvector presents unique challenges due to its interrelated components and the antipodal symmetry of the tensor. ^11,12^. Training on the two polar angles of the vector could result in two sets of θ and ∅, both viable solutions, potentially impeding model convergence. To address this, Cartesian coordinates of the tensor were used as the output of the model and the cost function was a cosine distance metric defined as:

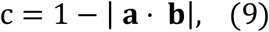

where **a** is the true vector and **b** is the vector predicted by the model. Since the loss function only focuses on direction, the model’s output must be normalized into a unit vector.

TensorFlow ^13^ was used to construct and train these models, leveraging its powerful capabilities in handling such neural network architectures and computations. The model structure is shown in Figure 3. The learning rate was 0.001 and training was conducted with the Adam optimiser ^14^. Each model was trained for 16 epochs.

**Figure 3.**
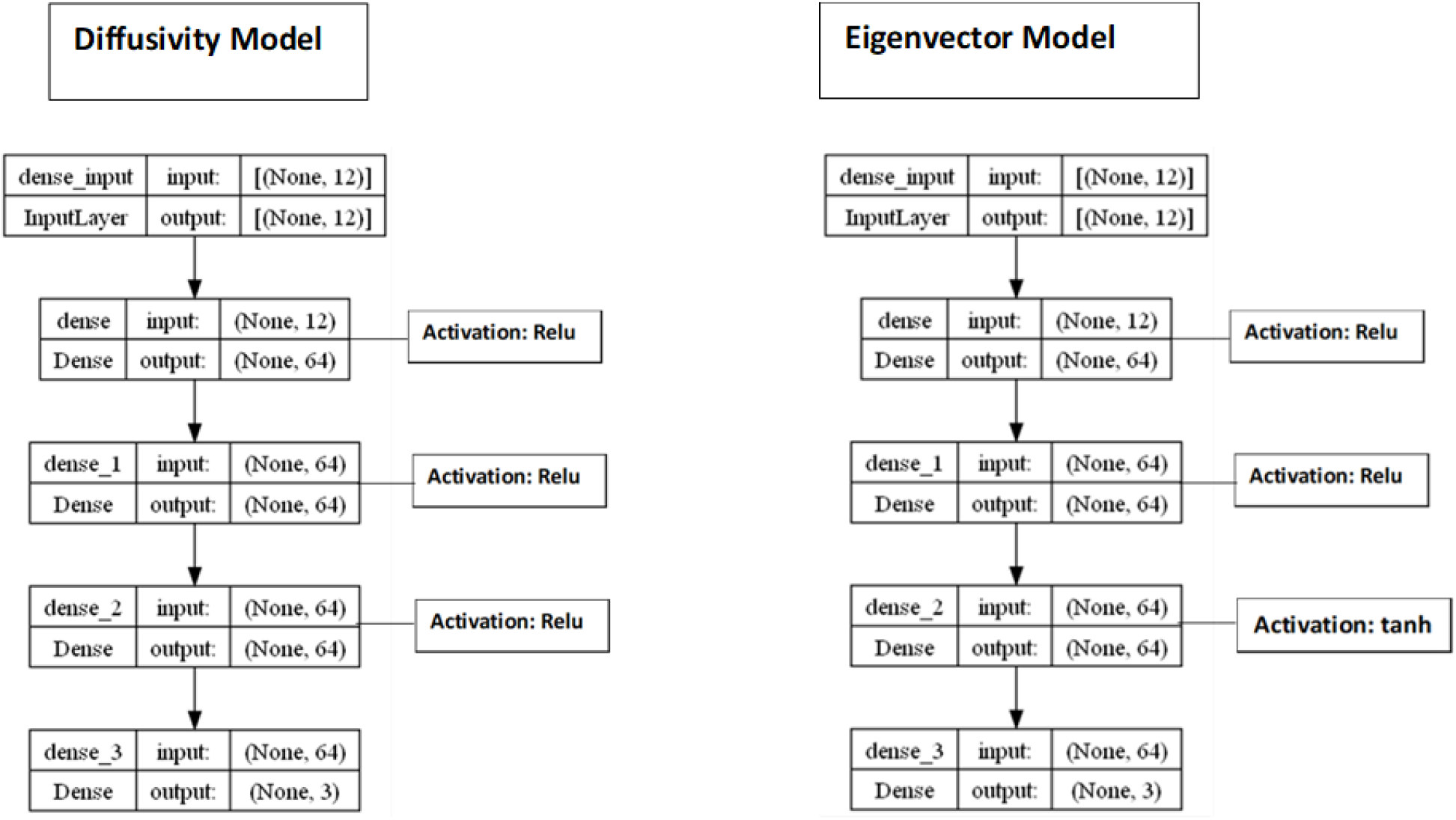
Graphical representation of the structure of the two models displayed through Tensorflow in Python. Each model is a multi-layer perceptron with three hidden layers between the input and output layers. A rectified linear unit (Relu) is used as the activation function for the hidden layers of the diffusivity model. The eigenvector uses the same, with a tanh activation in the last layer.

### 3.2 Tests

#### 3.2.1 Noise Susceptibility – Six directions vs four directions

We conducted a Monte Carlo simulation to compare noise effects on diffusivity estimation using 6 and 4 diffusion-encoding directions, respectively. Beginning with a diffusion tensor characterized with specified fractional anisotropy and eigenvalues, we set the trace of the tensor to 0.0021 mm^2^/s ^15,16^. Cylindrical symmetry was enforced, and we employed gradient vectors oriented along cube edges^17^, a b-value of 1000 s/mm^2^, and an arbitrary b_0_ signal value of XYZ. We generated six and four diffusion-weighted signals using the formula:

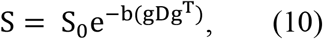

where 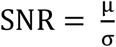, with μ representing the mean of the b0 signal and σ the standard deviation. Given a specific b0 signal and a predetermined SNR, we can simulate Rician noise in our signals ^9,15,16^. For each SNR, the eigenvalues were estimated using linear least squares for the six-direction approach. The standard tetrahedral method was used for the four-direction approach to estimate parallel and radial diffusivities. This was repeated for 2000 iterations.

#### 3.2.2 Noise Susceptibility – Standard Tetrahedral Approach vs Machine Learning Model

Using the same method, we generated four-directional signals for a diffusion tensor with a fractional anisotropy of 0.75 at varying SNR values. We estimated parallel and radial diffusivities using both the standard tetrahedral approach and our DL model. These tests employed a non-cylindrically symmetric tensor.

#### 3.2.3 Orientation Based Error

We tested whether our DL model could mitigate the errors in four-direction measurements when tensors are oriented close to the scanner’s axes. To do this, we generated non-cylindrically symmetric diffusion tensors with varying fractional anisotropy (FA) values and rotated each tensor by varying the polar angles (θ and ∅) of the principal eigenvector. Four-directional signals were generated for each orientation, and FA and tensor orientation were predicted using the standard tetrahedral approach and our DL model.

#### 3.2.4 Test on Images

The model was tested on a digital phantom from the Human Connectome Project, containing a DWI dataset with 128 gradient orientations ^18^, and on images taken at 64 mT from a single participant (male, 24 year-old) on a Hyperfine Swoop system. The study was approved by Cardiff University’s School of Psychology ethics committee. After providing written informed consent, the participant was instructed to remain still during the scan. Data were acquired using a diffusion-weighted fast spin echo (FSE) sequence ^19^ with tetrahedral encoding and a b-value of 900 s mm^-2^. Minimal post-processing was performed, limited to image co-registration using FSL’s FLIRT ^20^, without applying denoising. The goal was to compare the DL model’s performance against the standard tetrahedral approach in both digital phantom data and low field image data.

#### 3.2.5 Tensor Fitting and Tractography

Using the data taken at 64 mT, we generated a diffusion tensor for each voxel with the eigenvalues and principal eigenvectors estimated by our model. Then six diffusion-weighted images corresponding to six isotropically-distributed gradient orientations were synthetically generated from the tensors and the b0 image. This was done so that we could perform a DTI fit in software like MRtrix ^21^, which accepts a minimum of six diffusion-weighted images, while ensuring that the fitted tensors have the same anisotropy and main orientation as our model’s estimates for each voxel. Then, with the aid of a T1-weighted image, anatomically constrained tractography was attempted using MRtrix. The number of streamlines set was 1000000 and all other settings were left at their defaults.

## 4.0 Results

### 4.1 Noise Susceptibility – Six directions vs four directions

Figure 4 shows results from the six-direction linear least squares estimation, while Figure 5 shows the four-direction estimation using the standard tetrahedral approach. Due to significant differences in the magnitude of the eigenvalues, the parallel and radial diffusivities for the four-direction estimates were plotted separately. The four-direction estimates exhibit a greater deviation from true values at low SNR compared to the six-direction estimates, demonstrating the enhanced sensitivity of the standard tetrahedral approach to low SNR, and its limitations in low-field/low SNR datasets.

**Figure 4.**
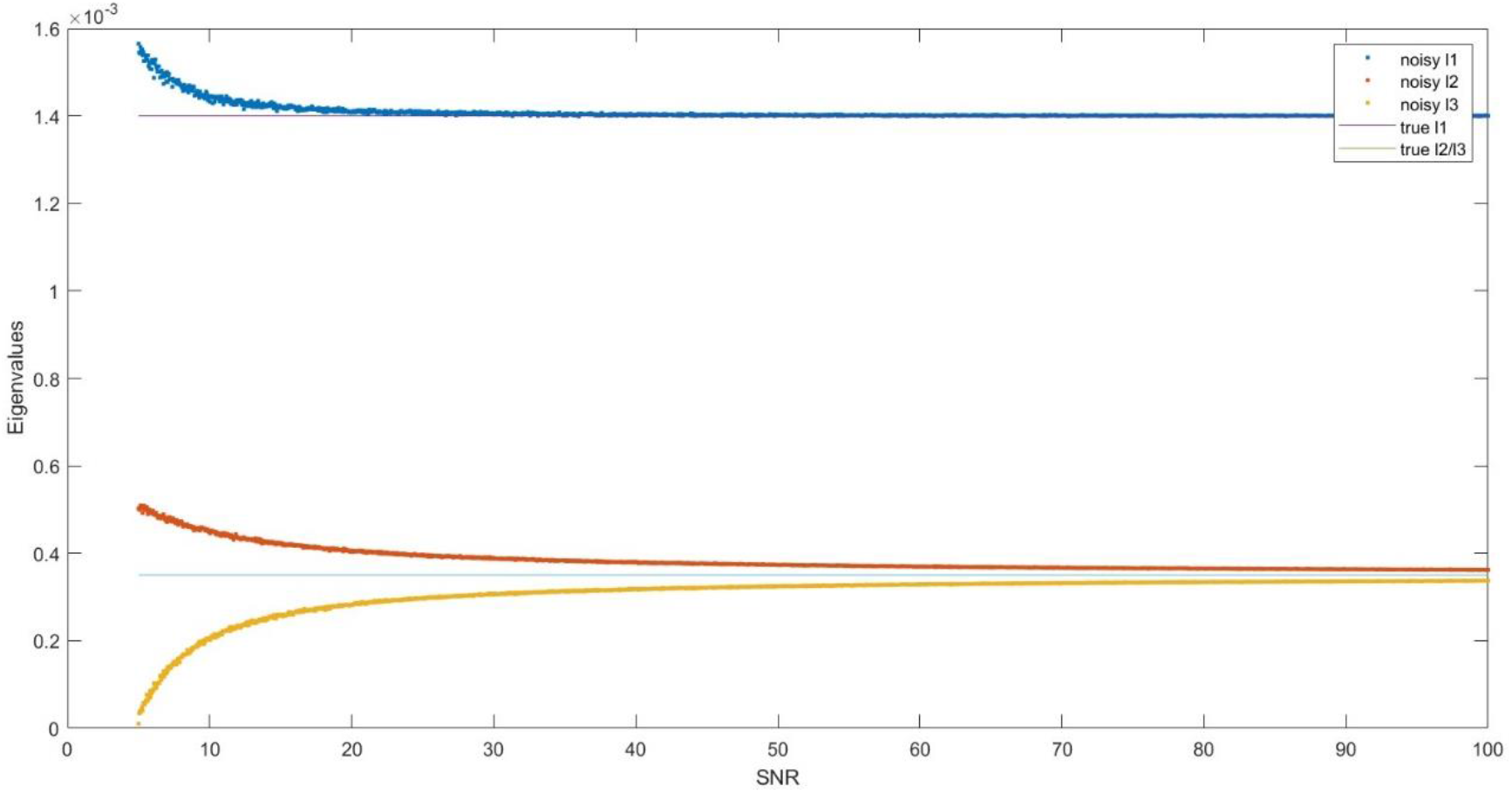
Effect of noise on the eigenvalue estimations (units = mm^2^/s) on a cylindrically symmetric tensor after a linear least squares estimation with 6 diffusion-weighted images. FA = 0.71, Trace = 0.0021 mm^2^/s.

**Figure 5.**
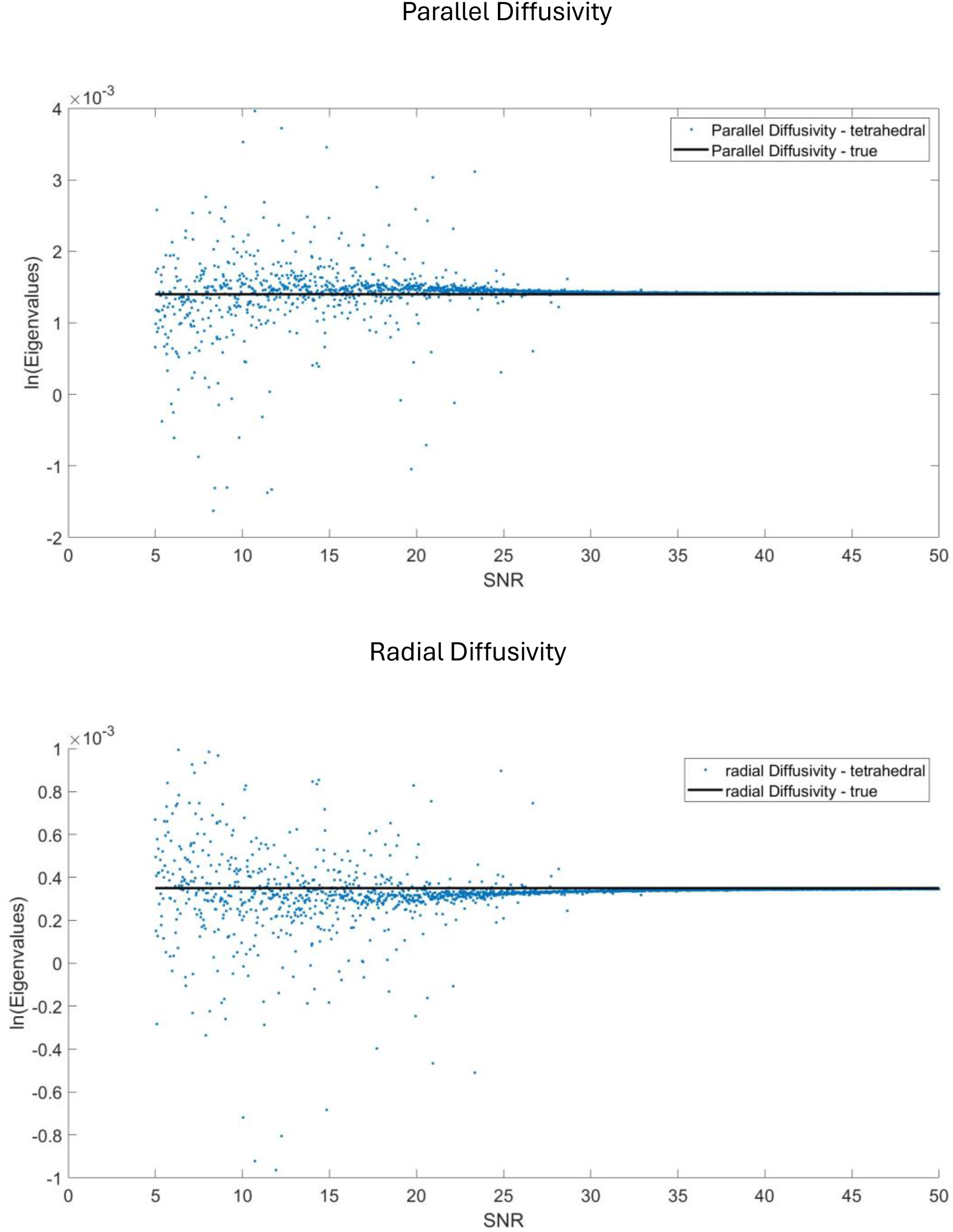
Effect of noise on the estimations of parallel and radial diffusivities (units = mm^2^/s) using the Conturo et al. tetrahedral method on a cylindrically symmetric tensor. FA = 0.71, Trace = 0.0021 mm^2^/s.

### 4.2 Noise Susceptibility – Standard Tetrahedral Approach vs DL Model

Figure 6 compares the predicted parallel and radial diffusivities from both the machine learning model and the standard tetrahedral approach across various SNR levels. The machine learning model shows greater stability and less deviation from the ground truth at low SNR compared to the standard tetrahedral approach, indicating better performance in noisy conditions.

### 4.3 Orientation Based Error

Figures 7 and 8 compare the machine learning model and the standard tetrahedral approach in predicting fractional anisotropy (FA) and fibre orientations for tensors oriented both close to and away from the scanner’s axes. Although errors persist, the machine learning model significantly reduces these errors compared to the standard tetrahedral approach, demonstrating higher accuracy in predictions.

### 4.4 Test on Images

Qualitative visual inspection of the digital phantom results in Figure 9 shows that the FA and orientation colour maps generated by the machine learning model more closely match the ground truth compared to those produced by the standard tetrahedral approach. In images taken at 64 mT (also shown in Figure 10), the standard tetrahedral approach produces incoherent FA maps, whereas the machine learning model highlights distinct tracts, indicating superior performance in accurately representing FA and fibre orientations. Figure 11 shows our preliminary attempts at performing tractography using this dataset.

## 5.0 Discussion

The noise susceptibility tests comparing six-direction linear least squares estimation and the standard tetrahedral approach reveal significant accuracy loss in diffusivity estimates at lower SNR values, especially when using four directions. This renders the standard tetrahedral approach unreliable for anisotropy estimation in low field scenarios with low SNR. However, the machine learning model mitigates the impact of SNR on four-direction diffusivity estimates. Although deviations from the ground truth persist, particularly at SNR values below 20, the ML-results are more coherent than those from the standard tetrahedral approach, showing a gradual continuous yet limited level deviation from the ground truth with each SNR, whereas the standard tetrahedral approach displays a wider and unpredictable level of deviation from the ground truth.

Figures 7 and 8 illustrate errors in the estimation of parallel and radial diffusivities and principal eigenvector orientation when tensors are aligned close to the scanner’s axes. While the errors are not eliminated entirely, they are significantly reduced. These tests used non-cylindrically symmetrical tensors, further highlighting the ML model’s superior performance over the standard tetrahedral approach in such cases. Armitage ^7^ has previously highlighted that for non-axisymmetric diffusion, the standard tetrahedral approach gives erroneous results while Lazar et al. ^22^ noted the impact of noise and partial volume effects on cylindrical symmetry. Despite imposing an assumption of cylindrical symmetry for tensor estimation, our method makes reliable predictions even when the measurements deviate from that assumption.

The digital phantom results in Figure 9 and 64 mT results in Figure 10 highlight the improvement in anisotropy and fibre orientation estimates. In the 64 mT data, white matter tracts are barely visible with the standard tetrahedral approach, whereas our method reveals tracts even in low SNR cases. While errors occurring when the tensor aligns with the scanner’s axes are reduced, they are not entirely removed, making our method unsuitable for advanced applications like whole-brain tractography. Future work could explore Bayesian approaches for uncertainty estimation or incorporating spatial priors to further reduce errors.

## 6.0 CONCLUSION

This work advances the approach of performing DT-MRI using only four diffusion-encoded measurements. Our solution overcomes the numerical instabilities that have hindered the robust application of earlier methods without introducing significant computational overhead or increased noise sensitivity. The implications of this advancement are particularly noteworthy, suggesting a pathway to significantly reduce DT-MRI scan durations, being especially valuable in ultra-low field (ULF) MRI applications, where time efficiency and noise management are critical concerns.

## Data Availability

All data produced in the present study are available upon reasonable request to the authors

https://github.com/jmametepe/tetrahedral_DTI

## Figures

**Figure 3.**
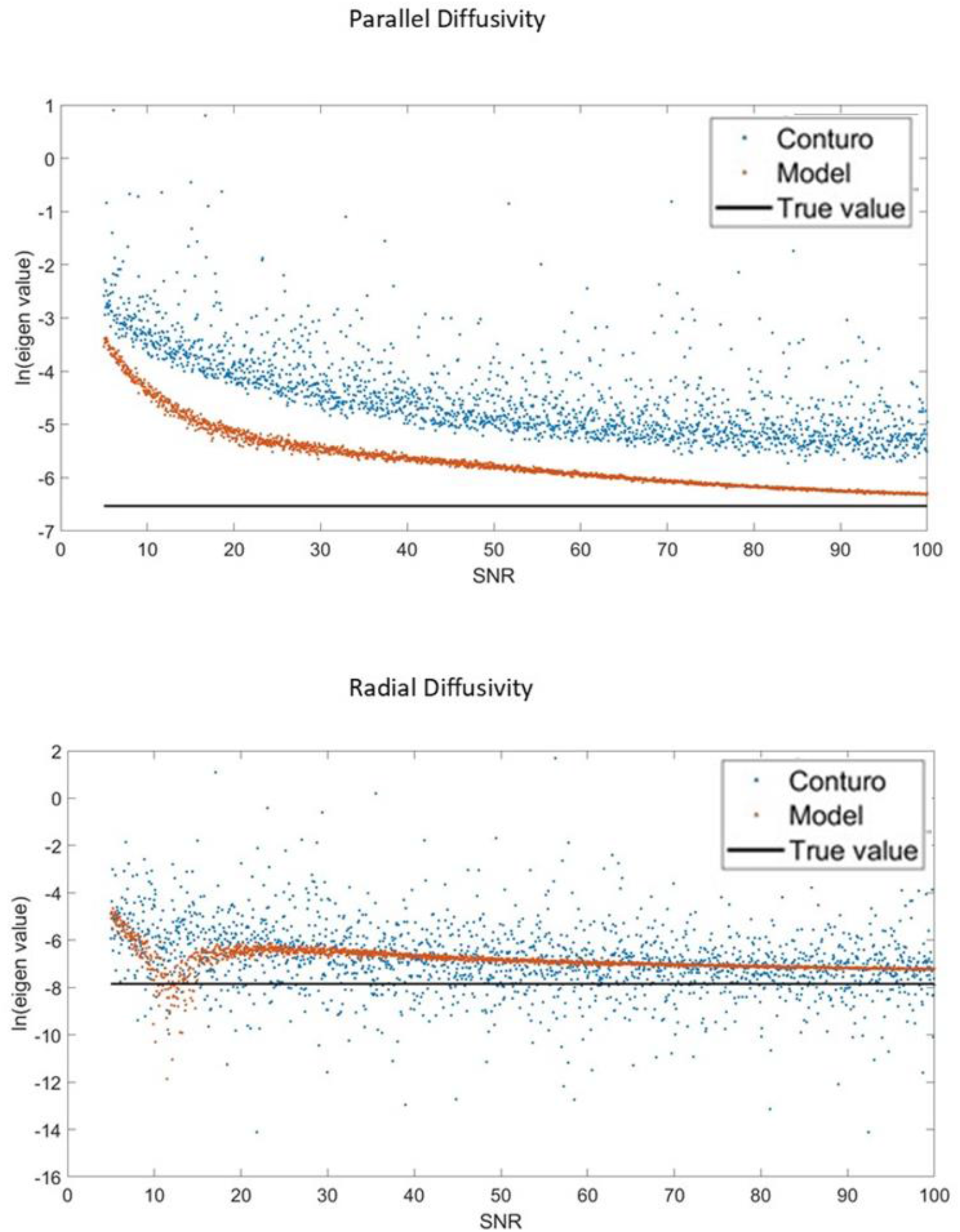
Comparison of the estimated parallel and radial diffusivities (units = mm^2^/s) of the model and the Conturo et al. tetrahedral approach with the true value on a natural log scale through a Monte Carlo Simulation. FA = 0.75, Trace = 0.0021 mm2/s. The model showed a greater robustness to noise as compared to the standard tetrahedral approach.

**Figure 4.**
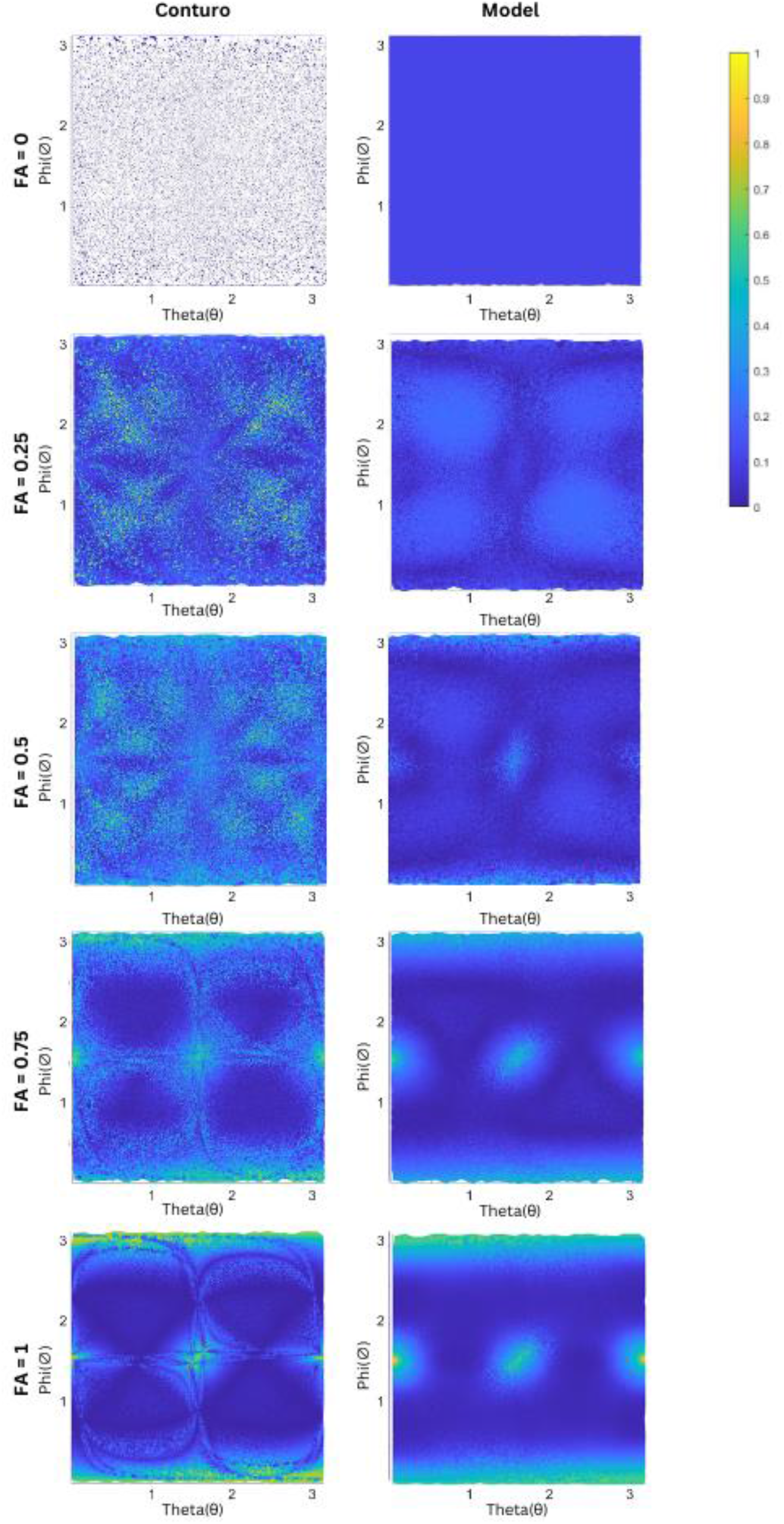
Absolute value of the FA error for various orientations of the principal eigenvector (θ, Φ). Trace = 0.0021 mm^2^/s. One of the gradient axes was rotated 12 degrees about the x-axis to simulate transformations due to registration.

**Figure 5.**
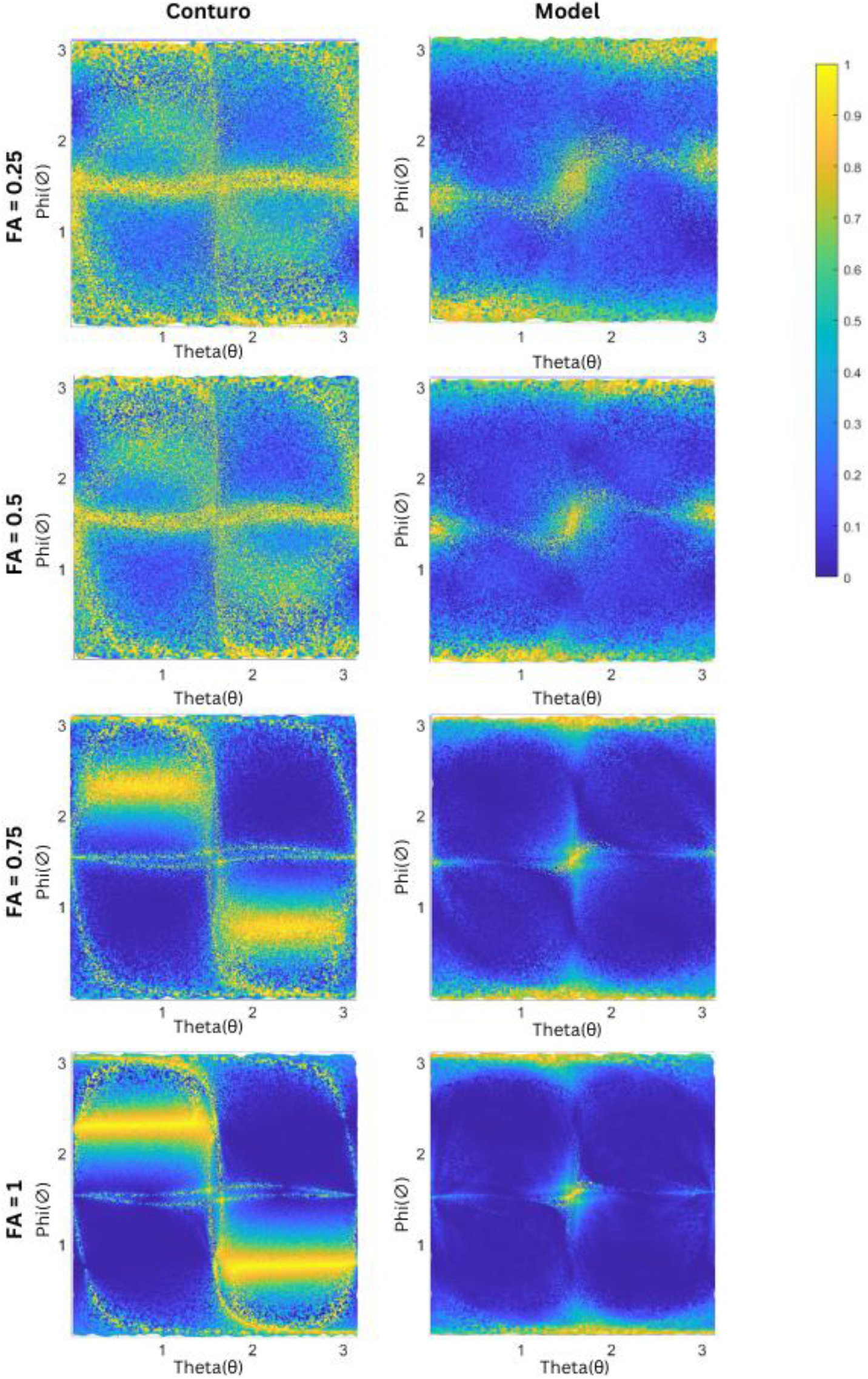
Angular error as the cosine distance between the predicted principal eigenvector and the true vector for various orientations of the vector (θ, Φ). Trace = 0.0021 mm^2^/s. One of the gradient axes was rotated 12 degrees in the x-axis to simulate transform

**Figure 6.**
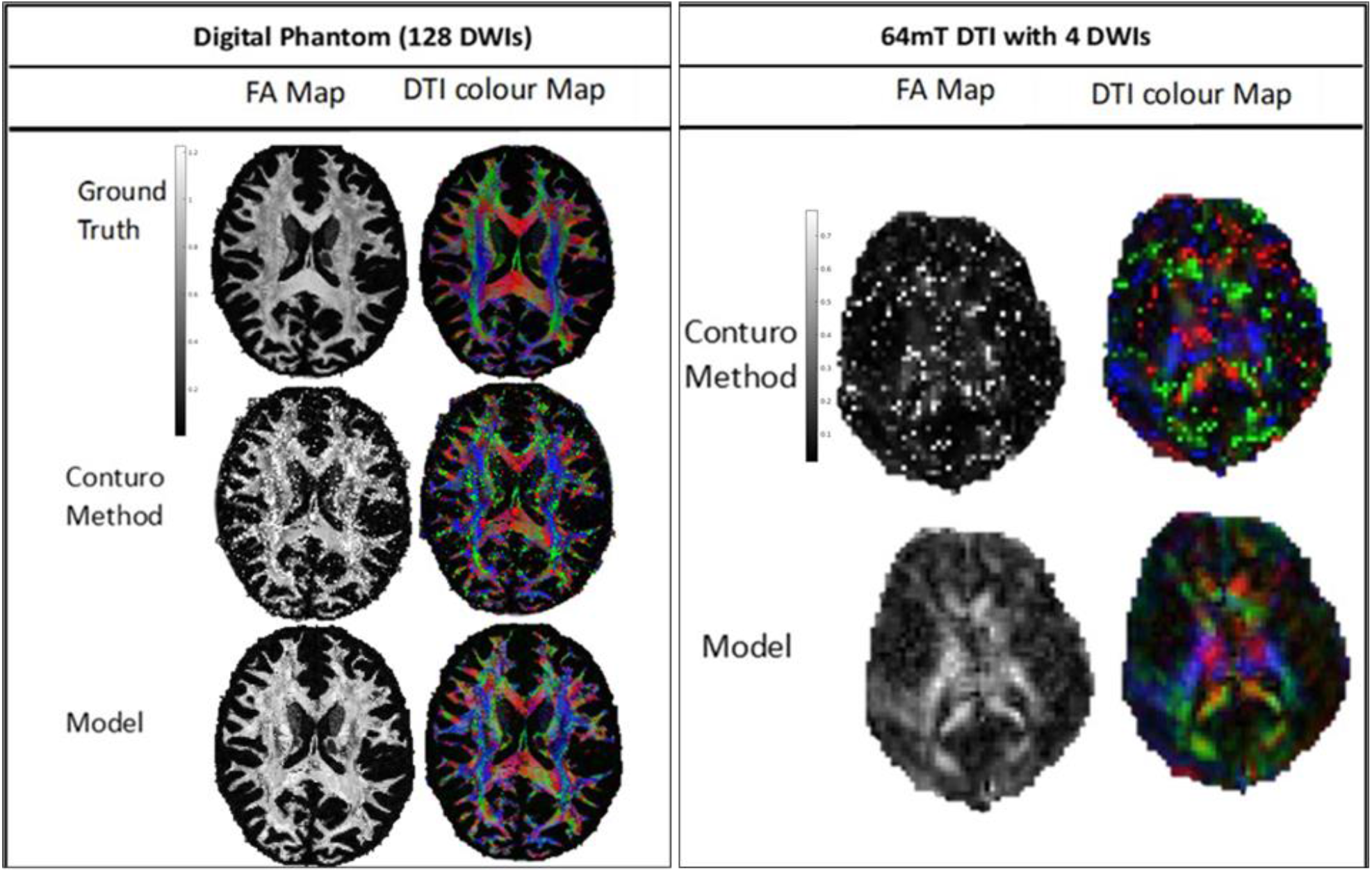
Results of XYZ using the model on the digital phantom and on images taken at 64 mT as compared with the Conturo et al. tetrahedral method.

**Figure 10.**
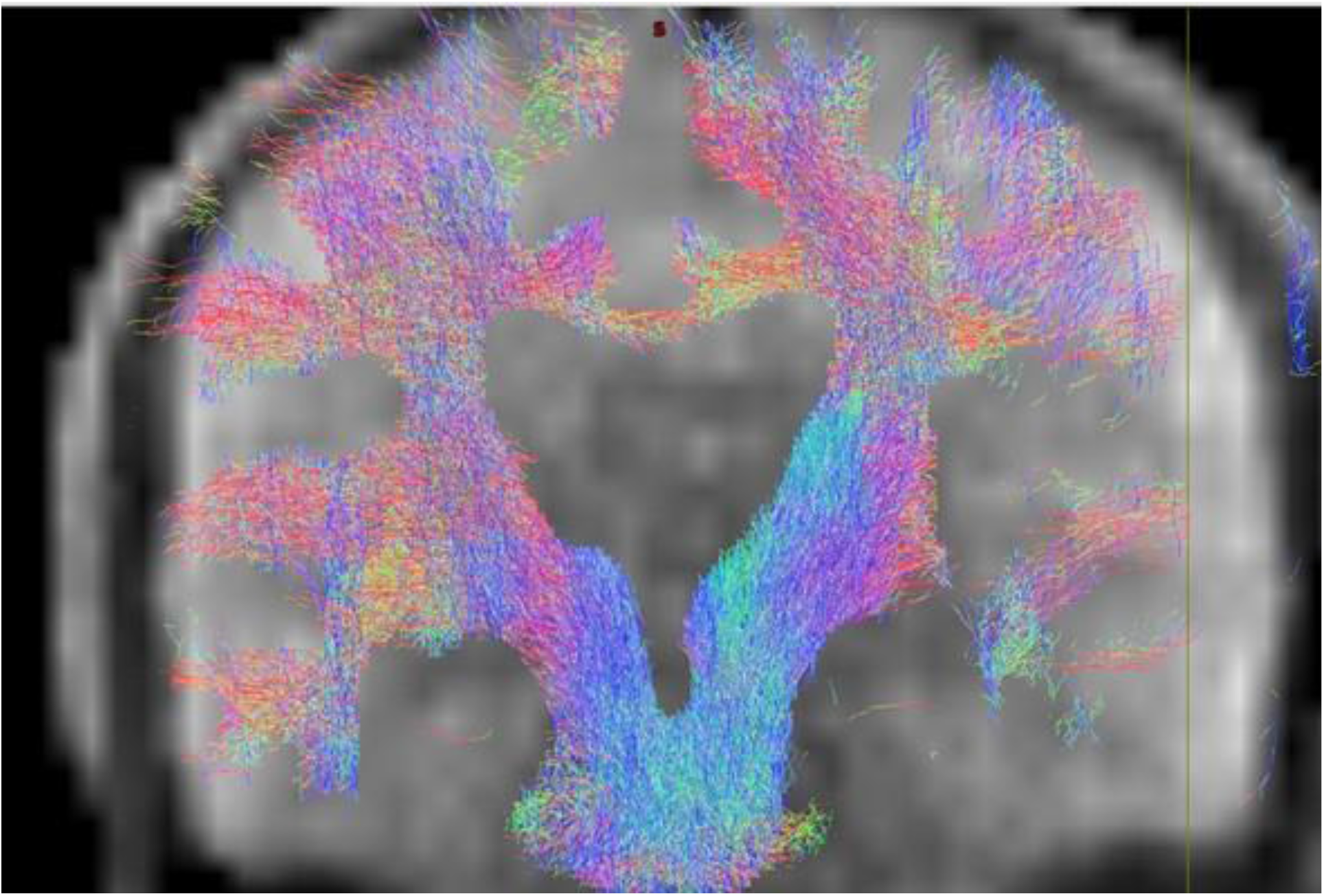
Preliminary attempts at anatomically constrained tractography using images taken at 64mT, with no denoising methods applied. The figure shows some unreliable results like fibres seemingly going through the skull. This further highlights the limitations of the model in advanced applications like tractography, particularly at low fields

## Data Availability

The code for the data generation and training of the model, as well as pretrained models can be found at https://github.com/jmametepe/tetrahedral_DTI

